# CSF PrP-amyloid seeding predicts clinical onset in inherited prion disease

**DOI:** 10.64898/2026.11.03.26367164

**Authors:** Tze How Mok, Akin Nihat, Sonia Minikel Vallabh, Nurit Omer, Noa Bregman, Michael Geschwind, Steven E. Arnold, Eric Vallabh Minikel, John Collinge, Simon Mead

**Author notes:** **Corresponding author**: Simon Mead.

## Abstract

Silent proteopathic seed propagation is hypothesised to precede neurodegenerative disease, but direct evidence that it signals near-term clinical conversion in humans is lacking. In prion disease, a prototypic disorder of proteopathic seeding, cerebrospinal fluid (CSF) real-time quaking-induced conversion (RT-QuIC) offers a direct measure of prion protein amyloid seeding activity. We analysed longitudinal CSF RT-QuIC data from 94 *PRNP* E200K mutation carriers across international natural history cohorts to determine whether seeding status predicts clinical conversion. Eleven carriers developed inherited prion disease; all showed at least one positive RT-QuIC result before onset. First positivity appeared a median of 2.0 years prior to symptoms (IQR 1.3–3.3; range 0.2–4.8). Landmark analyses showed RT-QuIC positivity identified a markedly increased short-term conversion risk, whereas persistently asymptomatic carriers remained negative throughout follow-up. These findings establish CSF RT-QuIC positivity as a biomarker-defined state of incipient conversion in E200K carriers, informing risk stratification and preventive trial design.

## MAIN TEXT

Prion disease is a rare but mechanistically distinctive neurodegenerative proteinopathy with a clearly defined therapeutic target: the host-encoded cellular prion protein (PrP). Pathogenesis is driven by propagation of disease-associated PrP assemblies, termed “prions”, through templated misfolding and recruitment of native PrP into pathogenic multimeric assemblies, culminating in neuronal dysfunction and death^1,2^ Most therapeutic efforts to date have involved repurposed drugs given after clinical onset, without convincing evidence of disease modification. By contrast, rationally designed PrP-targeting approaches, including monoclonal antibodies, antisense oligonucleotides and short-interfering RNA (siRNA), have strong preclinical support with some now in clinical evaluation.^3-6^ These strategies are most likely to succeed before established neurodegeneration, consistent with animal studies showing marked survival benefit when treatment is initiated in the preclinical phase.^6-9^

Prospective cohorts of asymptomatic carriers of prion protein gene (*PRNP*) mutations with a high lifetime risk of developing inherited prion disease (IPD) provide a potential setting for preventive clinical trials. However, such trials are constrained by low annual conversion rates and variable ages at onset, limiting statistical power.^10^ Biomarker-based enrichment strategies or alternative endpoints could improve trial efficiency. Early findings from UK and US cohorts demonstrated distinct biomarker trajectories in a handful of fast and slow converters. A small number of E200K mutation carriers were reported to show asymptomatic PrP-amyloid seeding activity in cerebrospinal fluid (CSF) by real-time quaking-induced conversion (RT-QuIC) for several years, followed by sharp rises in neurodegenerative markers and then clinical onset in quick succession, consistent with the two-phase kinetics of prion propagation first described in mouse models.^11,12^ Here, we assembled longitudinal datasets from PrionAtRisk Consortium members and applied statistical modelling to assess the predictive value of RT-QuIC status in E200K carriers.

We extracted longitudinal CSF RT-QuIC data from prospective natural history cohorts based in the US (Boston and San Francisco), UK (National Prion Monitoring Cohort) and Israel.^2,12,13^ A total of 94 individuals (55 females, 39 males; median age 57.5 (range 30 -86.6)) confirmed to carry the *PRNP* E200K mutation were identified, from whom 212 CSF samples were obtained (median follow-up 1.1 years, maximum 7.6 years). CSF samples from the Israeli cohort were tested using the first-generation RT-QuIC assay, while US and UK samples were tested using the second-generation RT-QuIC. Both are based on the same seeding assay principle, although the latter has faster kinetics^14^.

Of the 94 carriers, 11 developed Creutzfeldt-Jakob disease (CJD) during follow-up (8 females, 3 males; median age at conversion 62.1 (range 51.3–78.3)) (Fig. 1a) (Supplementary Table 1), while 83 remained asymptomatic (47 females, 36 males; median age at latest CSF sampling 54.5 (range 30-86.6)) (Fig. 1b) (Supplementary Table 2). All converters had at least one positive RT-QuIC result before clinical onset. In 4 of 11 converters, initial RT-QuIC samples were negative but became positive before onset (at 1.00, 1.56, 1.59 and 3.42 years before onset); in the remaining 7, the first available sample was already RT-QuIC positive (median 2.51 years before onset, range 0.18–4.79). Carriers who remained asymptomatic were RT-QuIC negative at all observed time points (median age at last sample 56.0 years, range 30.0–79.5). Repeated lumbar punctures were performed in 44 carriers (median interval 1.1 years; maximum follow-up 7.6 years); 23 had at least 3 years of follow-up and 8 had at least 5 years, and all remained RT-QuIC negative without clinical conversion.

**Figure 1:**
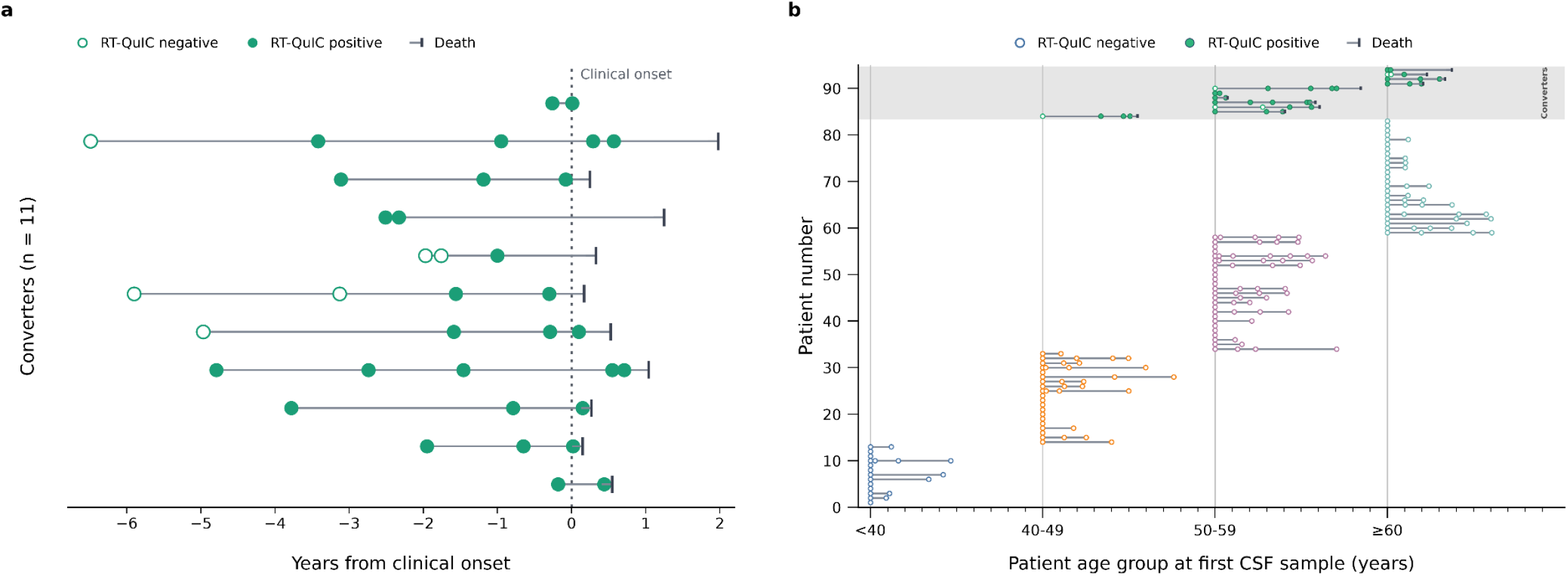
Longitudinal CSF RT-QuIC trajectories in E200K mutation carriers and converters. **a:** Serial CSF RT-QuIC measurements in E200K converters (n = 11), relative to clinical onset (anchored to year 0, dotted vertical line). Each horizontal line connects CSF samples (circles) for one subject; vertical ticks indicate death. **b:** Longitudinal CSF RT-QuIC measurements in converters and non-converters, grouped by age at first CSF sample. Each subject is represented by a horizontal trajectory, anchored to their age (decade) at first sample, and subsequent CSF samples are marked in years from this point (each minor tick reflects one year). The shaded region marks converters. In both panels a & b, unfilled circles denote RT-QuIC negativity, while filled circles denote RT-QuIC positivity; and vertical ticks indicate death.

RT-QuIC status showed marked prognostic separation. In a repeated landmark analysis, each CSF sample was treated as an independent landmark and classified according to RT-QuIC positivity and time to clinical onset (Fig. 2a). At 1, 2, 3 and 5 years after a landmark sample, the estimated proportion remaining asymptomatic was 57%, 33%, 19% and 0% for RT-QuIC-positive samples, compared with 100%, 97%, 97% and 89% for RT-QuIC-negative samples (Fig. 2a). A complementary subject-level analysis estimated risk of clinical conversion after subjects’ first positive or last classifiable negative RT-QuIC sample (Fig. 2b). This showed similarly high near-term risk after first positivity; the estimated proportion of subjects remaining asymptomatic after a first positive RT-QuIC sample was 73% at 1 year, 45% at 2 years and 36% at 3 years, compared with 100%, 96% and 96% after a last negative sample (Fig. 2b). Exploratory analyses did not show clear evidence that the predictive association between CSF RT-QuIC status and subsequent conversion was modified by age, sex or codon 129 genotype (Supplementary Table 3). Time from first RT-QuIC positivity to clinical onset was nominally longer in MV than MM carriers, but this comparison was based on small subgroup numbers (Supplementary Fig. 1).

**Figure 2:**
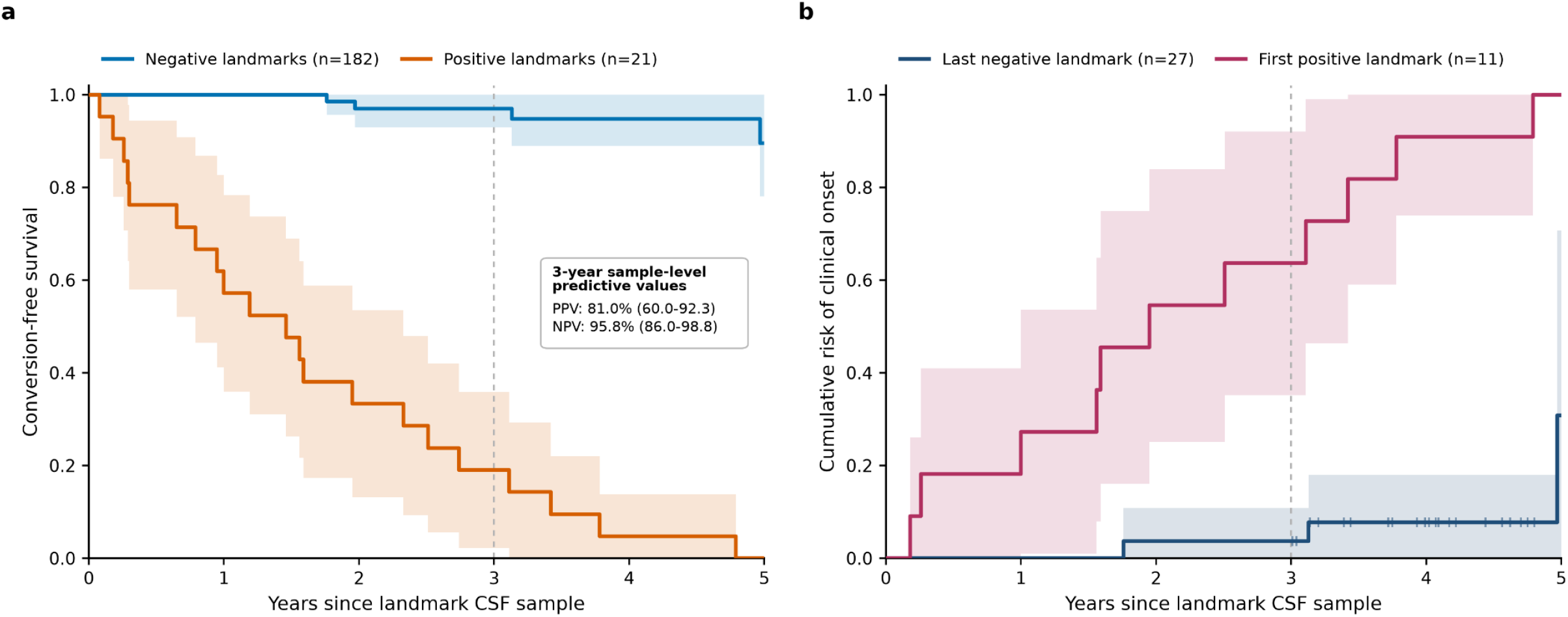
Longitudinal CSF RT-QuIC status and near-term risk of clinical onset in *PRNP* E200K mutation carriers. **a:** Repeated-landmark sample-level analysis. Each CSF sample was treated as a landmark and classified according to RT-QuIC status at the sampled time point (including multiple samples from the same patient). Displayed are Kaplan–Meier estimates of conversion-free survival from the time of sampling for RT-QuIC-positive and RT-QuIC-negative landmark samples, with 95% confidence intervals (Greenwood) shaded. The vertical dashed line marks the pre-specified 3-year prognostic horizon. Inset, Positive and Negative Predictive Values (PPV and NPV) for 3-year landmark-sample-level classification of 69 included CSF samples. **b:** Subject-level landmark analysis using first observed RT-QuIC-positive samples and last classifiable RT-QuIC-negative samples (minimum 3 years of follow-up or subsequent clinical conversion). Displayed are cumulative risks of clinical onset after the landmark, with shaded 95% confidence intervals. Tick marks indicate censoring after the subject-level landmark. Groups are not strictly mutually exclusive at the participant level, as some converters contributed an earlier negative landmark and a later first-positive landmark.

Using the repeated-landmark sample-level classification, RT-QuIC status identified clinical onset within 3 years of a given CSF sample with a sensitivity of 89.5% (95% CI 68.6, 97.1), specificity of 92.0% (95% CI 81.2, 96.8), positive predictive value of 81.0% (95% CI 60.0, 92.3) and negative predictive value of 95.8% (95% CI 86.0, 98.8) using 69 classifiable samples (17 true positives, 4 false positives, 46 true negatives and 2 false negatives – Fig. 2a, inset). This 3-year classification is conservative because it imposes a fixed prediction window on a biological process that may begin earlier. RT-QuIC positive samples obtained more than 3 years before onset are counted as false positives, even though they may represent early preclinical disease, while RT-QuIC-negative samples obtained before later conversion to positivity are counted as false negatives. Even under this stringent definition, prognostic accuracy remained high, with first positivity providing the clearest indication of near-term clinical onset.

Together, these findings define a clear sequence of biomarker progression and identify CSF RT-QuIC positivity as a strong state-transition marker of near-term clinical onset in *PRNP* E200K carriers. Within observed follow-up, carriers who had not converted remained RT-QuIC negative, arguing against persistent or intermittent non-consequential CSF PrP-amyloid seeding activity. Transition to RT-QuIC positivity appears to mark unidirectional entry into a clinically silent prodromal phase with sharply elevated short-term risk of onset. From a translational perspective, the observed interval from first positivity to onset suggests that annual sampling could provide a practical balance between detection of transition, procedural burden and opportunity for preventive intervention.

CSF RT-QuIC status also has immediate implications for risk counselling. Persistent negativity may provide reassurance over the short term, whereas first positivity identifies a substantially increased near-term risk of clinical onset within the observed time window. However, such information may also generate anxiety, anticipatory grief or misunderstanding about longer-term risk^15^. Any approach to disclosure should therefore be developed cautiously, with input from at-risk individuals, psychologists, clinical geneticists and ethicists, and offered only within a research framework with explicit informed consent, clear explanation of limitations, and integrated psychological and genetic counselling support.

Further, CSF RT-QuIC status has important implications for preventive trial design. RT-QuIC positivity identifies a preclinical state with substantially increased short-term risk of onset and may therefore support enrichment strategies or biomarker-informed endpoints. However, enrichment alone may not solve the practical problem of trial feasibility, because asymptomatic RT-QuIC-positive carriers are uncommon and may be slow to identify and recruit. These data also point to at least two distinct biological transitions: from RT-QuIC negativity to RT-QuIC positivity, and from RT-QuIC positivity to clinical onset. Trials should prospectively incorporate RT-QuIC in order to assess the timing of these transitions; further work is needed to determine whether quantitative measures derived from the assay can capture longitudinal change within the prodromal phase.

Finally, although based on a modest number of converters, these E200K RT-QuIC data provide unusually strong evidence that a binary seeding biomarker can mark a clinically meaningful state transition preceding onset in human neurodegeneration. This should encourage development of bespoke or more widely applicable CSF RT-QuIC assays for other major inherited prion diseases, including P102L, D178N and octapeptide repeat insertions,^16^ as well as evaluation of peripherally accessible tissues and biofluids that might augment or eventually reduce reliance on CSF sampling.^17-20^ More broadly, these findings support a translational paradigm relevant to other neurodegenerative proteinopathies, particularly amyloid beta, tau and α-synuclein pathologies, in which detection of an early proteopathic seeding transition could improve risk stratification, disease monitoring and the design of preventive trials.

## Data Availability

All data produced in the present study are available upon reasonable request to the author, subject to respective data transfer agreements.
All data produced in the present work are contained in the manuscript

## Ethics

The UK National Prion Monitoring Cohort was approved by the Scotland A Research Ethics Committee (REC code 05/MRE00/63). The Boston study was approved by the Mass General Brigham IRB (2017P000214). The San Francisco study (TLC-Pre-gPrD) was apprived by the University of California San Francisco IRB. The Israeli study was approved by the Tel Aviv Medical Center IRB and the Israeli Ministry of Health IRB (0484-17 and 0215-18).

