## Supplementary Material for "CSF PrP-amyloid seeding predicts clinical onset in inherited prion disease"

### Supplementary materials

#### Supplementary Table 1 CSF RT-QuIC status of E200K converters

| Converter | Sex | Codon 129 | Age range at 1st CSF (years) | RT-QuIC Status | Time to onset (years) |
|---|---|---|---|---|---|
| **1** | F | MM | 50-59 | Negative | -5.9 |
| | | | | Negative | -3.13 |
| | | | | Positive | -1.56 |
| | | | | Positive | -0.30 |
| | | | | Death | 0.17 |
| **2** | F | MM | 50-59 | Positive | -3.78 |
| | | | | Positive | -0.79 |
| | | | | Positive | 0.15 |
| | | | | Death | 0.27 |
| **3** | F | MV | 50-59 | Positive | -4.79 |
| | | | | Positive | -2.74 |
| | | | | Positive | -1.46 |
| | | | | Positive | 0.55 |
| | | | | Positive | 0.71 |
| | | | | Death | 1.04 |
| **4** | F | MM | 50-59 | Positive | -0.18 |
| | | | | Positive | 0.44 |
| | | | | Death | 0.55 |
| **5** | M | MM | > 60 | Positive | -1.95 |
| | | | | Positive | -0.65 |
| | | | | Positive | 0.02 |
| | | | | Death | 0.15 |
| **6** | M | MV | 40-49 | Negative | -4.97 |
| | | | | Positive | -1.59 |
| | | | | Positive | -0.29 |
| | | | | Positive | 0.10 |
| | | | | Death | 0.53 |
| **7** | F | VV | > 60 | Negative | -1.97 |
| | | | | Negative | -1.76 |
| | | | | Positive | -1.00 |
| | | | | Death | 0.33 |
| **8** | F | MV | > 60 | Positive | -2.51 |
| | | | | Positive | -2.33 |
| | | | | Death | 1.25 |
| **9** | M | MV | > 60 | Positive | -3.11 |
| | | | | Positive | -1.19 |
| | | | | Positive | -0.08 |
| | | | | Death | 0.25 |
| **10** | F | MM | 50-59 | Positive | -0.26 |
| | | | | Positive | 0.01 |
| **11** | F | MV | 50-59 | Negative | -6.49 |
| | | | | Positive | -3.42 |
| | | | | Positive | -0.95 |
| | | | | Positive | 0.29 |
| | | | | Positive | 0.57 |
| | | | | Death | 1.98 |

Supplementary Table 2 CSF RT-QuIC status of asymptomatic E200K mutation carriers

| Carrier | Sex | Codon 129 | Age Range at 1st CSF (years) | RT-QuIC status | Time from 1st CSF (years) |
|---|---|---|---|---|---|
| **1** | F | MM | < 40 | Negative | 0 |
| **2** | F | MM | > 60 | Negative | 0 |
| | F | MM | > 60 | Negative | 4.62 |
| **3** | F | MV | 40-49 | Negative | 0 |
| | F | MV | 40-49 | Negative | 3.99 |
| **4** | M | MV | < 40 | Negative | 0 |
| | M | MV | < 40 | Negative | 1.11 |
| **5** | M | MV | < 40 | Negative | 0 |
| **6** | M | MM | > 60 | Negative | 0 |
| | M | MM | > 60 | Negative | 1.55 |
| | M | MM | > 60 | Negative | 2.49 |
| | M | MM | > 60 | Negative | 3.72 |
| **7** | F | MM | > 60 | Negative | 0 |
| | F | MM | > 60 | Negative | 4.01 |
| | F | MM | > 60 | Negative | 6.02 |
| **8** | M | MM | > 60 | Negative | 0 |
| | M | MM | > 60 | Negative | 1.97 |
| | M | MM | > 60 | Negative | 4.98 |
| | M | MM | > 60 | Negative | 6.06 |
| **9** | M | MM | < 40 | Negative | 0 |
| | M | MM | < 40 | Negative | 0.92 |
| **10** | M | MM | 50-59 | Negative | 0 |
| | M | MM | 50-59 | Negative | 1.31 |
| | M | MM | 50-59 | Negative | 2.36 |
| | M | MM | 50-59 | Negative | 7.06 |
| **11** | F | MV | 40-49 | Negative | 0 |
| | F | MV | 40-49 | Negative | 1.11 |
| | F | MV | 40-49 | Negative | 2.38 |
| **12** | M | MV | > 60 | Negative | 0 |
| | M | MV | > 60 | Negative | 0.98 |
| | M | MV | > 60 | Negative | 4.15 |
| | M | MV | > 60 | Negative | 5.73 |
| **13** | F | MM | 50-59 | Negative | 0 |
| | F | MM | 50-59 | Negative | 0.31 |
| | F | MM | 50-59 | Negative | 2.32 |
| | F | MM | 50-59 | Negative | 3.69 |
| | F | MM | 50-59 | Negative | 4.87 |
| **14** | F | MM | 40-49 | Negative | 0 |
| | F | MM | 40-49 | Negative | 0.18 |
| | F | MM | 40-49 | Negative | 1.53 |
| | F | MM | 40-49 | Negative | 5.97 |
| **15** | M | MM | 50-59 | Negative | 0 |

| | | | | | |
|---|---|---|---|---|---|
| | M | MM | 50-59 | Negative | 0.23 |
| | M | MM | 50-59 | Negative | 1.05 |
| | M | MM | 50-59 | Negative | 3.21 |
| | M | MM | 50-59 | Negative | 4.36 |
| | M | MM | 50-59 | Negative | 5.36 |
| | M | MM | 50-59 | Negative | 6.41 |
| **16** | F | MM | 40-49 | Negative | 0 |
| **17** | F | MV | 40-49 | Negative | 0 |
| | F | MV | 40-49 | Negative | 0.21 |
| | F | MV | 40-49 | Negative | 0.97 |
| | F | MV | 40-49 | Negative | 4.99 |
| **18** | F | VV | 40-49 | Negative | 0 |
| | F | VV | 40-49 | Negative | 1.26 |
| | F | VV | 40-49 | Negative | 2.3 |
| **19** | F | VV | < 40 | Negative | 0 |
| | F | VV | < 40 | Negative | 1.21 |
| **20** | F | MV | > 60 | Negative | 0 |
| **21** | F | MM | 50-59 | Negative | 0 |
| **22** | F | MV | > 60 | Negative | 0 |
| | F | MV | > 60 | Negative | 1.21 |
| **23** | M | MM | < 40 | Negative | 0 |
| **24** | M | MM | 50-59 | Negative | 0 |
| | M | MM | 50-59 | Negative | 0.23 |
| | M | MM | 50-59 | Negative | 2.81 |
| | M | MM | 50-59 | Negative | 3.93 |
| | M | MM | 50-59 | Negative | 5.65 |
| **25** | M | MV | 40-49 | Negative | 0 |
| | M | MV | 40-49 | Negative | 1.97 |
| | M | MV | 40-49 | Negative | 4.07 |
| | M | MV | 40-49 | Negative | 4.98 |
| **26** | F | MM | 40-49 | Negative | 0 |
| **27** | F | MV | < 40 | Negative | 0 |
| | F | MV | < 40 | Negative | 0.27 |
| | F | MV | < 40 | Negative | 1.62 |
| | F | MV | < 40 | Negative | 4.66 |
| **28** | F | MM | > 60 | Negative | 0 |
| **29** | F | MV | 50-59 | Negative | 0 |
| | F | MV | 50-59 | Negative | 1.09 |
| | F | MV | 50-59 | Negative | 2.02 |
| **30** | F | MM | 50-59 | Negative | 10.69 |
| **31** | M | MM | 50-59 | Negative | 0 |
| | M | MM | 50-59 | Negative | 1.2 |
| | M | MM | 50-59 | Negative | 2.57 |
| | M | MM | 50-59 | Negative | 4.17 |
| **32** | M | MM | 50-59 | Negative | 0 |

| | | | | | |
|---|---|---|---|---|---|
| **33** | M | MV | > 60 | Negative | 0 |
| | M | MV | > 60 | Negative | 1.12 |
| | M | MV | > 60 | Negative | 2.42 |
| **34** | M | MM | 50-59 | Negative | 0 |
| | M | MM | 50-59 | Negative | 1.45 |
| | M | MM | 50-59 | Negative | 2.47 |
| | M | MM | 50-59 | Negative | 4.07 |
| **35** | F | MM | > 60 | Negative | 0 |
| | F | MM | > 60 | Negative | 1.04 |
| | F | MM | > 60 | Negative | 2.02 |
| | F | MM | > 60 | Negative | 3.75 |
| **36** | F | MM | > 60 | Negative | 0 |
| | F | MM | > 60 | Negative | 1.04 |
| **37** | F | MM | > 60 | Negative | 0 |
| | F | MM | > 60 | Negative | 1.2 |
| **38** | M | MM | 50-59 | Negative | 0 |
| | M | MM | 50-59 | Negative | 1.12 |
| | M | MM | 50-59 | Negative | 2.61 |
| | M | MM | 50-59 | Negative | 4.26 |
| **39** | M | MM | > 60 | Negative | 0 |
| **40** | M | MM | > 60 | Negative | 0 |
| **41** | F | MM | 50-59 | Negative | 0 |
| **42** | F | MM | 40-49 | Negative | 0 |
| | F | MM | 40-49 | Negative | 1.24 |
| | F | MM | 40-49 | Negative | 2.52 |
| **43** | F | MM | 50-59 | Negative | 0 |
| | F | MM | 50-59 | Negative | 1.56 |
| **44** | F | MM | 40-49 | Negative | 0 |
| | F | MM | 40-49 | Negative | 1.79 |
| **45** | M | MM | 40-49 | Negative | 0 |
| **46** | F | MM | > 60 | Negative | 0 |
| **47** | M | MV | 40-49 | Negative | 0 |
| **48** | F | MM | 50-59 | Negative | 0 |
| **49** | F | MM | 50-59 | Negative | 0 |
| **50** | M | MM | 50-59 | Negative | 0 |
| | M | MM | 50-59 | Negative | 1.47 |
| | M | MM | 50-59 | Negative | 2.98 |
| **51** | F | MM | 50-59 | Negative | 0 |
| | F | MM | 50-59 | Negative | 1.17 |
| **52** | M | MM | 40-49 | Negative | 0 |
| **53** | F | MM | 40-49 | Negative | 0 |
| **54** | F | MM | 50-59 | Negative | 0 |
| | F | MM | 50-59 | Negative | 2.13 |
| **55** | M | MV | 50-59 | Negative | 0 |
| **56** | M | MM | > 60 | Negative | 0 |

| | | | | | |
|---|---|---|---|---|---|
| **57** | F | MM | 50-59 | Negative | 0 |
| **58** | M | MM | 50-59 | Negative | 0 |
| **59** | F | MM | 50-59 | Negative | 0 |
| **60** | M | MV | > 60 | Negative | 0 |
| | M | MV | > 60 | Negative | 1 |
| | M | MV | > 60 | Negative | 2.09 |
| **61** | F | MV | > 60 | Negative | 0 |
| | F | MV | > 60 | Negative | 1.03 |
| **62** | M | VV | 40-49 | Negative | 0 |
| | M | VV | 40-49 | Negative | 1.2 |
| | M | VV | 40-49 | Negative | 2.13 |
| **63** | F | MM | 50-59 | Negative | 0 |
| | F | MM | 50-59 | Negative | 2.59 |
| | F | MM | 50-59 | Negative | 3.58 |
| | F | MM | 50-59 | Negative | 4.8 |
| **64** | M | MV | 40-49 | Negative | 0 |
| | M | MV | 40-49 | Negative | 4.17 |
| | M | MV | 40-49 | Negative | 7.6 |
| 65 | F | MV | > 60 | Negative | 0 |
| 66 | F | MV | > 60 | Negative | 0 |
| 67 | M | MM | > 60 | Negative | 0 |
| 68 | M | MV | < 40 | Negative | 0 |
| 69 | M | MM | 50-59 | Negative | 0 |
| | M | MM | 50-59 | Negative | 1.02 |
| | M | MM | 50-59 | Negative | 3.33 |
| | M | MM | 50-59 | Negative | 4.94 |
| 70 | M | MM | < 40 | Negative | 0 |
| | M | MM | < 40 | Negative | 4.22 |
| 71 | M | MV | 40-49 | Negative | 0 |
| 72 | M | MM | > 60 | Negative | 0 |
| 73 | F | MV | < 40 | Negative | 0 |
| | F | MV | < 40 | Negative | 3.39 |
| 74 | F | MM | < 40 | Negative | 0 |
| 75 | M | MM | < 40 | Negative | 0 |
| 76 | F | MM | > 60 | Negative | 0 |
| 77 | M | MM | 40-49 | Negative | 0 |
| 78 | F | MV | 40-49 | Negative | 0 |
| | F | MV | 50-59 | Negative | 1.05 |
| 79 | F | MV | 40-49 | Negative | 0 |
| 80 | F | MV | 50-59 | Negative | 0 |
| 81 | F | MM | 50-59 | Negative | 0 |
| 82 | F | MM | < 40 | Negative | 0 |
| 83 | F | M/V | > 60 | Negative | 0 |
| | F | M/V | > 60 | Negative | 1.06 |

Supplementary Table 3 Cox Proportional Hazards Analysis

| Covariate | Hazard ratio | 95% CI | P value |
|---|---|---|---|
| **Age (per 10 years)** | 1.65 | 0.44 – 6.19 | 0.46 |
| **Male sex** | 2.10 | 0.37 – 12.13 | 0.41 |
| **Codon 129 MV vs MM** | 0.32 | 0.03 – 3.04 | 0.32 |
| **Codon 129 VV vs MM** | 4.06 | 0.07 – 226.05 | 0.49 |
| | **Likelihood Ratio** | | **P value** |
| **Overall site effect** | 2.89 | | 0.41 |

We undertook a Cox proportional hazards model, fitted using time from first RT-QuIC positive CSF sample to clinical onset as the outcome, to assess whether the association between RT-QuIC positivity and subsequent clinical conversion were independent of demographic and cohort-related factors. Covariates included age (per 10-year increase), sex, *PRNP* codon 129 genotype, and recruitment site. Adjustment was performed by including these variables simultaneously in the model, allowing estimation of the effect of each factor while holding the others constant. No significant associations were observed for age, sex, codon 129 genotype, or site. These findings suggest that the observed prognostic value of RT-QuIC positivity is not explained by differences in age, sex, genotype, or site.

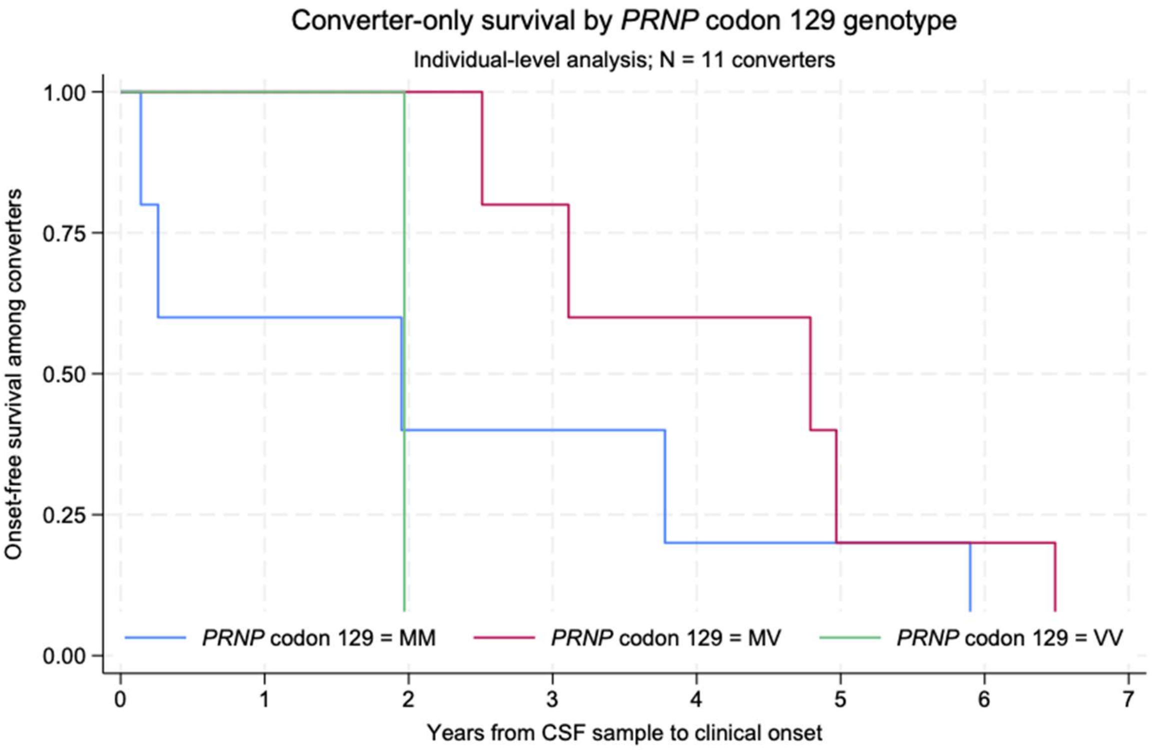

**Supplementary Figure 1** Kaplan–Meier estimates of onset-free survival following first RT-QuIC-positive CSF sample among the 11 converters, stratified by *PRNP* codon 129 genotype (MM, n=5; MV, n=5; VV, n=1).
